# Culture-free whole genome sequencing of *Mycobacterium tuberculosis* using ligand-mediated bead enrichment method

**DOI:** 10.1101/2024.01.23.24301340

**Authors:** Shruthi Vasanthaiah, Renu Verma, Ajay Kumar, Aravind Bandari, John George, Mona Rastogi, Gowrang Kasaba Manjunath, Jyoti Sharma, Abhishek Kumar, Janavi Subramani, Kiran Chawla, Akhilesh Pandey

## Abstract

**Background:** Direct whole genome sequencing (WGS) of *Mycobacterium tuberculosis* (*Mtb*) can be used as a tool to study drug resistance, mixed infections, and within host diversity. However, WGS is challenging from clinical samples due to low number of bacilli against a high background.

**Methods:** We prospectively collected 34 samples (sputum, n=17; bronchoalveolar lavage, BAL, n=13 and pus, n=4) from patients with active tuberculosis (TB). Prior to DNA extraction, we used a ligand-mediated magnetic bead method to enrich *Mtb* from clinical samples and performed WGS on Illumina platform.

**Results:** *Mtb* was definitively identified based on WGS from 88.2% (30/34) of the samples of which 35.3% (12/34) were smear negative. The overall median genome coverage was 15.2% (IQR = 7.9-39.3). There was a positive correlation between load of bacilli on smears and genome coverage (p-value < 0.001). We detected 58 genes listed in the WHO mutation catalogue in each positive sample (median coverage = 85%, IQR = 61%-94%), enabling the identification of mutations missed by routine diagnostics. Mutations causing resistance to rifampicin, isoniazid, streptomycin, and ethambutol were detected in 5/34 (14.7%) samples, including the *rpoB* S441A mutation that confers resistance to rifampicin which is not covered by Xpert MTB/RIF. This approach also allowed us to identify mixed infections in eight samples (BAL=4/8, pus=2/3 and sputum= 2/10) including samples that were infected with three or more different strains of *Mtb*.

**Conclusions:** We demonstrate the feasibility of magnetic bead-based enrichment for culture-free WGS of *Mtb* from clinical specimens, including smear-negative samples. This approach can also be integrated with low-cost sequencing workflows such as targeted sequencing for rapid detection of *Mtb* and drug resistance.

## Introduction

Tuberculosis remains a major public health problem. As per the recent WHO report, there was a 3.9 % increase in TB incidence rate between 2020 - 2022 (1). India contributes to the world’s highest TB incidence, with approximately 3 million cases annually (2). Over the last decade, due to the accessibility of low-cost benchtop sequencers, streamlined workflows, and a faster turnaround time, whole genome sequencing has rapidly evolved from a mere research tool to a clinical platform with the ability to identify the pathogen and its resistance to drugs (3), investigate transmission (4), and conduct public health surveillance (5–7). However, whole-genome sequencing has not yet entered the mainstream of clinical diagnosis in tuberculosis, mainly due to the requirement of culturing *Mtb* prior to sequencing. (7–9).

Unlike most laboratory procedures, culture based *Mtb* analysis remains challenging due to slow growth of mycobacteria, laborious procedures, and expensive biocontainment facility requirement precluding its use in resource limited settings (10). Additionally, in culture-based assays, typically a few single colony subcultures are taken for the subsequent molecular analysis and sequencing. This may create a bias due to under recognition of heterogeneity and mixed infections (8, 11–13). Rapid molecular assays such as Xpert MTB/RIF (Cepheid, Sunnyvale, CA, USA), GenoType MTBDRplus and MTBDRsl assays (Hain Life science GmbH, Germany) detect *Mtb* directly from clinical samples with a shorter turnaround time (14). However, these assays are limited by the number of mutations they can target that can lead to false negatives, and do not provide information to investigate the pathogen genome (15).

Utilizing *Mtb* DNA isolated directly from clinical samples can not only be used as a rapid diagnostic tool, genomic analysis of *Mtb* from clinical samples may also provide valuable insights into lineage prevalence, within host diversity and evolution of drug resistance.

However, WGS of *Mtb* from direct samples is challenging due to low bacterial load and poor recovery of *Mtb* during sample processing (9,16). In a metagenomic analysis, Doughty and colleagues provided the first proof of principle on WGS of *Mtb* from sputum samples without enrichment. They sequenced eight positive sputum samples to detect *Mtb* and perform metagenomic analysis. However, despite high sputum bacillary load, the sequencing coverage achieved was very low and the samples could not be used for basic genomic analysis such as detection of resistance, mixed infection and identifying novel mutations (17). Brown and colleagues subsequently used biotinylated RNA baits to capture the *Mtb* DNA extracted from sputum samples, and accurately detected drug-resistant phenotypes (18).

Using RNA baits, Goig et al. further sequenced *Mtb* directly from sputum samples and delineated the transmission clusters of tuberculosis (19). Verma et al. further used RNA baits to sequence *Mtb* from pooled surface swabs collected from prison cells occupied by patients with active TB to study transmission (20). While these studies demonstrated the importance of sequencing the *Mtb* genome without culture, due to high cost, long genome capture protocol (∼ 48hours), and requirement of highly trained personnel, capture assays are less scalable in resource-constrained settings. Thus, further studies are required to explore cost- effective and alternate enrichment methods that can be employed for WGS of *Mtb* directly from clinical samples.

Ligand-coated magnetic bead technology which specifically enriches *Mtb* from clinical samples has been previously used for concentrating *Mtb* for fluorescent microscopy and culture. It was found that magnetic bead-based methods significantly increased the sensitivity of microscopy and positivity of *Mtb* on culture (21,22). However, this bead based *Mtb* enrichment method for whole genome sequencing of clinical samples has not been explored yet. Also, studies on WGS of *Mtb* from other sample types than sputum have been lacking, and the sensitivity of enrichment in these samples is yet to be explored. This is especially important in the case of extrapulmonary TB infections and samples from pediatric patients where bacillary load in the sputum samples is either undetectable or absent (23–25).

In this study, we used a ligand-mediated magnetic bead strategy to enrich *Mtb* from sputum, BAL, and Pus samples from patients with active TB, and performed WGS. We further compared our findings with routinely performed diagnostic tests including smear microscopy, Xpert MTB/RIF, and culture. We assessed the utility of further exploring this enrichment method in culture-free rapid diagnosis of *Mtb*, detection of drug resistance, mixed infections, and phylogeny.

## Methods

### Patient enrollment and ethical statement

We enrolled 34 patients with confirmed active tuberculosis cases at the Kasturba medical college, Manipal, India between June to September 2018. All participants were aged 18 years or older and provided written consent prior to study initiation. The study was approved by the institutional review board of Kasturba medical college, Manipal Academy of Higher Education (IEC:526/2018).

### Sample collection

We collected approximately 1ml clinical specimen from active TB patients in replicates. This included early morning sputum (n=17), bronchoalveolar lavage (BAL; n=13) and pus (n=4). BAL and pus samples were collected aseptically. One tube was used for the routine microbiological testing and reporting, and the other tube was used for magnetic bead enrichment and DNA extraction for sequencing. All samples were screened for *Mtb* bacilli using Zeihl-Neelsen staining and classified as scanty, 1+, 2+, and 3+ based on the Revised National Tuberculosis Control Program (RNTCP) guidelines (26). The samples were also tested on Xpert/MTB RIF (Cepheid, Sunnyvale, USA) assay and inoculated on liquid (Mycobacterial growth indicator test, MGIT) and solid culture (Lowenson-Jenson, LJ) media. The *Mtb* growth was monitored for 45 days on MGIT and eight weeks on LJ slants. Five sputum samples which were negative for *Mtb* on microscopy, Xpert/MTB RIF and culture were taken as negative controls and enriched with magnetic beads followed by DNA extraction.

### Sample processing

The samples collected were processed using TB-Bead kit (Microsens Diagnostics Ltd, London, UK) following the manufacturer’s instructions in a BSL3 facility. Briefly, the samples were thinned by adding equal volume of thinning solution and allowed to stand at room temperature for 20 minutes after mixing. The homogenized samples were mixed with equal volume of magnetic beads conjugated with the ligand. After mixing gently, the samples and the bead slurry were allowed to stand at room temperature for 5 minutes to capture the mycobacterial cells. The slurry was then placed on a magnetic stand to separate, and supernatant was discarded. The beads were washed with 10 mM NaOH in quick succession prior to the elution of the bound mycobacteria from the beads using a 100ul elution buffer.

Eluate containing enriched mycobacteria was used for DNA extractions. Workflow schema for bead enrichment and WGS is shown in **Figure 1**.

**Figure 1:**
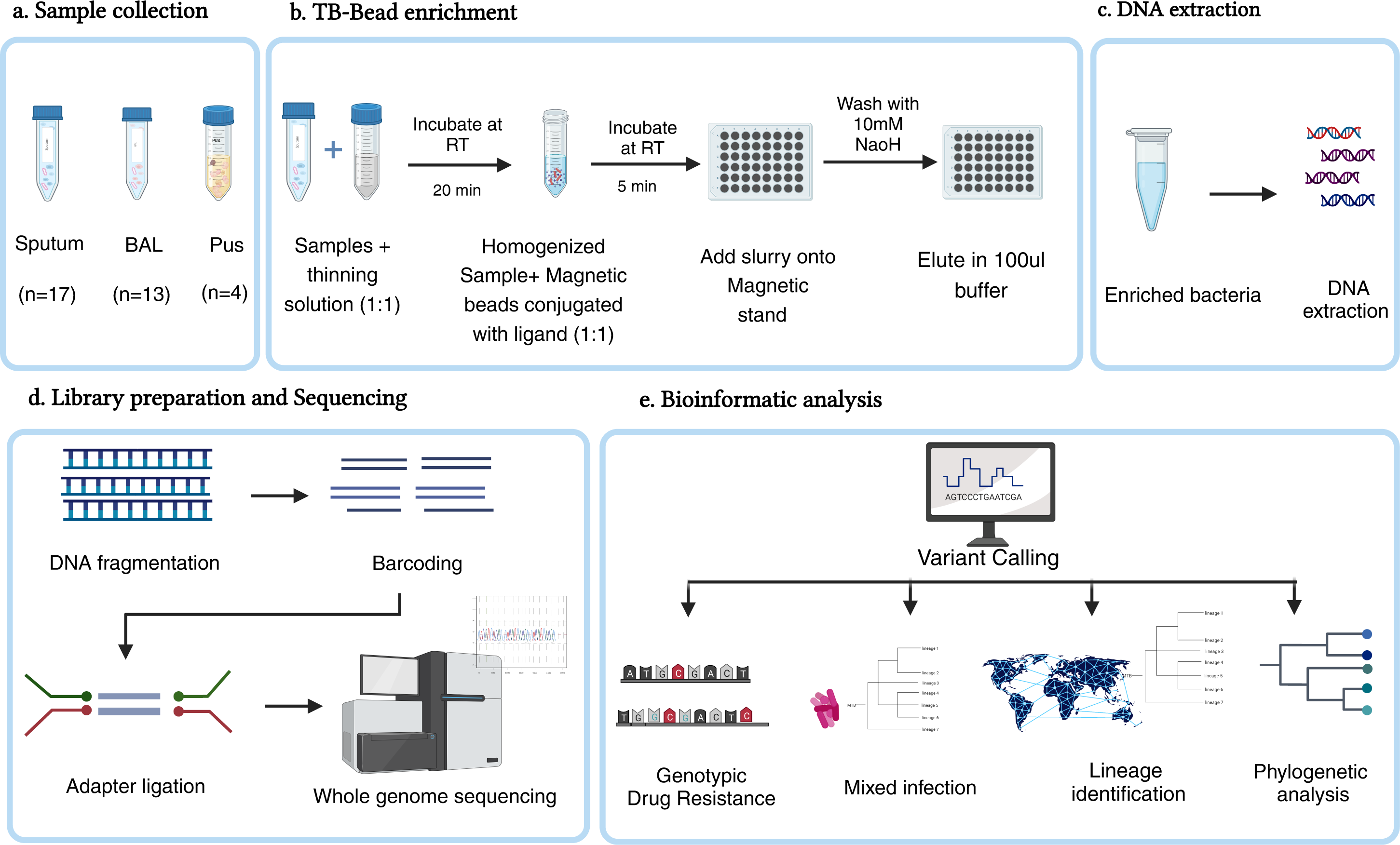
Workflow schema for bead based *Mtb* enrichment and WGS: For bead concentration, sample is mixed with equal volume of thinning solution and homogenized after incubation. Slurry is mixed with magnetic beads conjugated with ligands to enrich *Mtb* cells and incubated for binding. The *Mtb* cells are eluted in 100ul of elution buffer which is subsequently used for DNA extraction. The extracted DNA is quality tested and used for library preparation using Nextera XT DNA Library Preparation Kit, Illumina. Bioinformatic analysis is performed to detect resistance, phylogeny, mixed infections, and new mutations.

#### 2.3.1 DNA extraction and library preparation

The eluate enriched for mycobacteria was homogenized using the MP Biomedical bead beater (MP Biomedicals) and DNA was extracted using QIAamp DNA Mini Kit, according to the manufacturer’s instructions. The DNA eluted from the column in 10mM Tris buffer was stored at -80 **°**C until further use. Isolated genomic DNA was quantified using qubit fluorometric method. Indexed libraries were prepared for each of the samples using a low-input genomic DNA library preparation kit (Nextera XT DNA Library Preparation Kit, Illumina) as per the manufacturer’s instructions. Briefly, DNA isolated from the samples was fragmented and the DNA fragments obtained were subjected to end-repair before the ligation of indexed adapter sequences. The indexed libraries were further amplified using PCR to obtain the required amount of indexed library from each sample considering the low DNA yield. Qualities of the libraries were assessed using Agilent Bio Analyzer before proceeding with the clustering and sequencing process. The clustered libraries were sequenced on Illumina HiSeq XTen platform to obtain 150 bp paired-end reads.

#### 2.3.2 Whole genome sequencing and data acquisition

About 0.5GB data was generated per sample using the high output mode, to provide a depth of about 100X for the microbial genome. Quality of the acquired data in FastQ file formats was assessed using FastQC (v0.11.5) and processing of the raw reads was carried out using FastQC fqtrim (v0.9.4) (27) tools. Quality processed reads were mapped against ‘Mycobacteria’ (Taxonomy ID: 1763) sequences using Bowtie2 (v2.2.5) (28) . To rule out the DNA contamination from possible host organisms, we aligned raw reads against the reference genome of human (*Homo sapiens*), build 39, downloaded from NCBI. To identify the pathogenic bacteria from the whole genome sequencing data, we aligned the processed reads against representative bacterial/archaeal genomes database available at NCBI, using Bowtie2 with aforementioned parameters. This database comprised of genome sequences from 4,378 bacterial species, including various Mycobacterial species.

Quality of the raw sequencing data was checked using FastQC. Quality trimming and adapter removal was carried out using AdapterRemoval (Version: 2.1.7) (29). After performing quality control, the raw sequencing data was aligned against the complete genome of Mycobacterium tuberculosis H37Rv (RefSeq Accession: NC_000962.3) using BWA-sampe using default parameters (30). PCR duplicates were removed and the reads mapping to the reference genome were fetched and counted using Samtools (https://github.com/samtools/). A total of 336 complete genomes of different mycobacterium species were downloaded from NCBI, that includes the available complete genomes of different strains of *Mycobacterium tuberculosis*. All mapped reads were aligned to these complete mycobacterium genomes using blastn program and the reads that were matching with 100% identity were fetched. Further, reads that are unique to H37Rv were fetched using a python program.

### Variant calling, drug resistance and phylogenetic analysis

Variant calling was performed using of bcftools (Version 1.18.) (31). We used *TBProfiler* (32) tool to carry out the identification of mutations associated with anti-TB drug resistance.

We compared the list of mutations derived from *TBProfiler* with the comprehensive list provided by the WHO for the association of drug resistance (33). We also performed phylogenetic analysis of enriched samples using *TBProfiler* to classify samples into lineages and sub lineages and detect mixed infections. We used the MAFFT tool (Version 7) to generate multiple sequence alignment files and constructed a phylogenetic tree using RAxML-NG (Version 0.9.0).

### Statistical analysis

We performed Kruskal-Wallis test to determine if there is a significant difference in the genome coverage based on smear grading. Samples were divided into negative, scanty, 1+, 2+ and 3+ based on smear grading, and p values were calculated. We also performed Kruskal-Wallis test to determine if the percentage of reads that mapped to the H37Rv genome varied based on the sample type. All analyses were performed in R (Version 4.3.1).

## Results

### Patient characteristics

A total 34 individuals with confirmed active TB were recruited for the study. Among these, 73.5% (25/34) of the participants were male (median age= 49 years; IQR= 33.5). Median age of the female participants was 39 years (IQR=21.5). The samples consisted of sputum (n=17), BAL (n=13) and Pus (n=4). Out of 34 samples, 64.7% were positive on smear microscopy.

Among those, the samples were AFB negative (n=10), scanty bacilli (n=4), AFB 1+ (n=7), AFB2+ (n=6) and AFB 3+ (n=3). A total of 76.5% of the samples were positive for *Mtb* growth on MGIT and LJ culture, and 91.2% samples were positive on Xpert MTB/RIF. We did not observe any *Mtb* growth on culture or amplification in PCR for negative controls. (**Table 1**).

**Table 1:** Clinical and microbiological characteristics of the study cohort.

| **Clinical characteristic** | **Total (n=34)\*** | **BAL(n=13)** | **PUS(n=4)** | **Sputum(n=17)** |
| --- | --- | --- | --- | --- |
| No of patients (no. [%]) | 34(100) | 13(38.2) | 4(11.8) | 17(50) |
| Male (no. [%]) | 25(73.5) | 7(53.8) | 2(50) | 16(94.1) |
| Median (IQR) age (yr) | 44.5(28–57) | 44(27-50) | 39(27-61.5) | 55(35.5-63) |
| **No. (%) of patients with smear grade at baseline** |  |  |  |  |
| 3+ | 3(8.8) | 2(15.4) | 0(0) | 1(5.9) |
| 2+ | 6(17.6) | 1(7.7) | 0(0) | 5(29.4) |
| 1+ | 8(23.5) | 0(0) | 0(0) | 8(47) |
| Scanty | 5(14.7) | 2(15.4) | 1(25) | 2(11.8) |
| Negative | 12(35.3) | 8(61.5) | 3(75) | 1(5.9) |
| **GeneXpert/PCR** |  |  |  |  |
| Positive | 31(91.2) | 13(100) | 4(100) | 14(82.4) |
| Negative | 3(8.8) | 0(0) | 0(0) | 3(17.6) |
| **Culture (MGIT/LJ)** |  |  |  |  |
| Positive | 26(76.5) | 8(61.5) | 1(25) | 17(100) |
| Negative | 8(23.5) | 5(38.5) | 3(75) | 0(0) |

\* Microbiologically positive Samples

### Whole genome sequencing of bead enriched *Mtb*

*Mtb* reads were detected in the majority of the samples (30/34; 88.2%) with 35.3% (12/34) samples being smear negative. Total number of *Mtb*-specific reads detected in 30 samples which passed the quality cutoff was 38,31,740 (median reads per sample= 80, 673). The samples had a median percentage genome coverage 15.2 % (IQR= 7.9-31.4). There was a significant difference in percentage of H37Rv genome coverage among samples with high vs low smear positivity (Kruskal Wallis p-value< 0.001) **Figure 2A**. Sequencing statistics based on various sample types is provided in **Table 2**.

**Figure 2:**
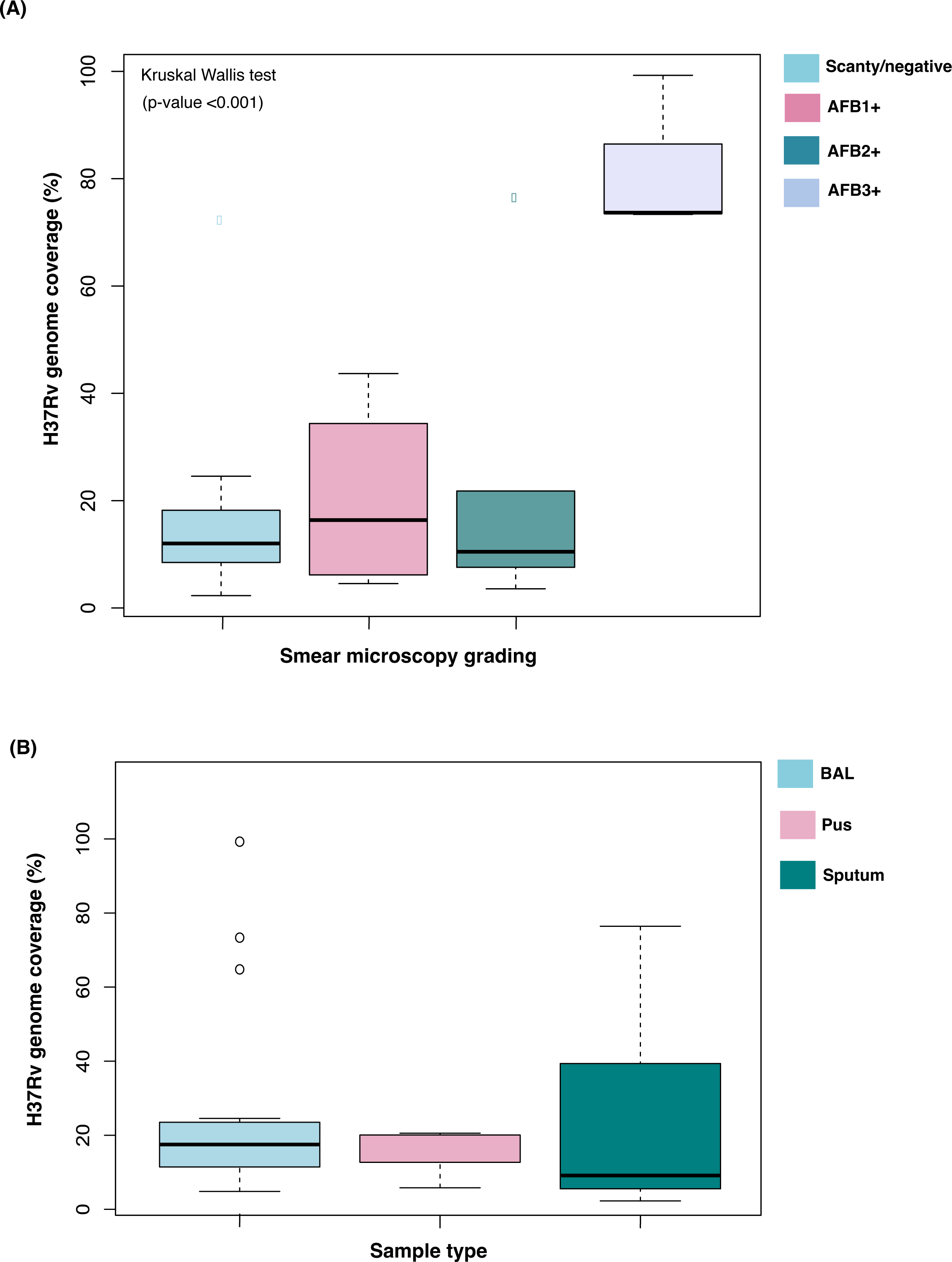
(A) H37Rv genome coverage vs smear microscopy grading: There was a significant difference in percentage of H37Rv genome coverage among samples with high vs low smear positivity (p-value< 0.001). **(B) H37Rv genome coverage (%) vs sample type:** The median H37Rv genome coverage in BAL, pus and sputum was 10.5% 15.4% and 19.6% respectively. Out of total number of reads, the percentage of reads mapped to H37Rv genome were significantly low in sputum samples when compared with BAL and Pus.

**Table 2:** Sequencing statistics for the quality filtered samples enriched using ligand-based magnetic beads.

| Category | Sample type |  |  |
| --- | --- | --- | --- |
|  | BAL (n=11) | Sputum (n=16) | Pus (n=3) |
|  | (Median, IQR) | (Median, IQR) | (Median, IQR) |
| Total reads (n)* | 104,638 (125,002) | 54,557 (51,994) | 187,140 (104,874) |
| Mean baseQ | 39.5 (39.5) | 38.9 (39) | 39.1 (38.9) |
| Reads mapped to H37Rv (n) | 114,978 (138,590) | 57,996 (54,427) | 205,084 (120,696) |
| H37Rv genome coverage (%)<sup>\$</sup> | 15.4 (99.2-6.8) | 10.5 (76.4-2.3) | 19.6, (19.6-5.8) |
| H37Rv genome coverage depth (x) <sup>\$</sup> | 0.63 (18.7-0.2) | 0.6, (5.1-0.1) | 1.1 (1.1-0.6) |

\*Total reads after quality filtering; <sup>\$</sup> Maximum and minimum values provided in the bracket

Despite low bacillary count as per AFB staining, BAL and pus samples had higher *Mtb* genome coverage than sputum samples. Number of smear negative samples in sputum, BAL and pus were 1/17, 8/13 and 3/4, respectively. The median H37Rv genome coverage in sputum, BAL and pus was 10.5% 15.4% and 19.6%, respectively **Figure 2B**. Out of total number of reads, the percentage of reads mapped to H37Rv genome were significantly low in sputum samples when compared with BAL and pus (p-value= 0.0147) (**Supplementary figure 1a and b**). It has been previously observed that pus samples due to low *Mtb* load are difficult to culture for subsequent analysis (34). This makes it challenging to analyze these samples on a comprehensive DST panel or perform any genomic analysis. We observed that due to less complexity of BAL and pus samples, the overall bead enrichment yield was higher for these samples allowing whole genome sequencing.

### Drug resistance detection

We compared variant calling data obtained from whole-genome sequencing (WGS) of enriched samples with fifty-eight genes from *Mtb* genome listed by the WHO as genes associated with drug resistance (33). We detected reads mapping to these genes in all the samples with median percentage coverage 85% (IQR= 61%-94%) (**Supplementary Table 1a**).

We detected mutations in one or more genes in twenty-four samples from the list of fifty-eight genes (**Supplementary Table 1b**). A total one hundred and eighty-nine mis-sense mutations were detected in nine genes. These mutations were categorized as non-synonymous (n=114) and mutations in the non-coding exon (n=75). The significance of most of these mutations need to be studied further. Among all the resistance-associated genes analyzed, we detected mutations in five genes (*rpoB, katG, rrs, inhA and embB*) that confer resistance to first and second-line anti-TB drugs in 22/30 (73.3%) samples. The H37Rv percentage genome coverage is shown in (**Figure 3A**) S450L mutation in *rpoB* gene that confers high-level resistance to rifampicin was detected in one BAL sample previously detected as RIF resistant on Xpert MTB/RIF. In addition to confirming RIF resistance, we also detected mutation in *katG* (Ser315Thr) and inhA (Ser94Ala) conferring resistance to Isoniazid.

**Figure 3:**
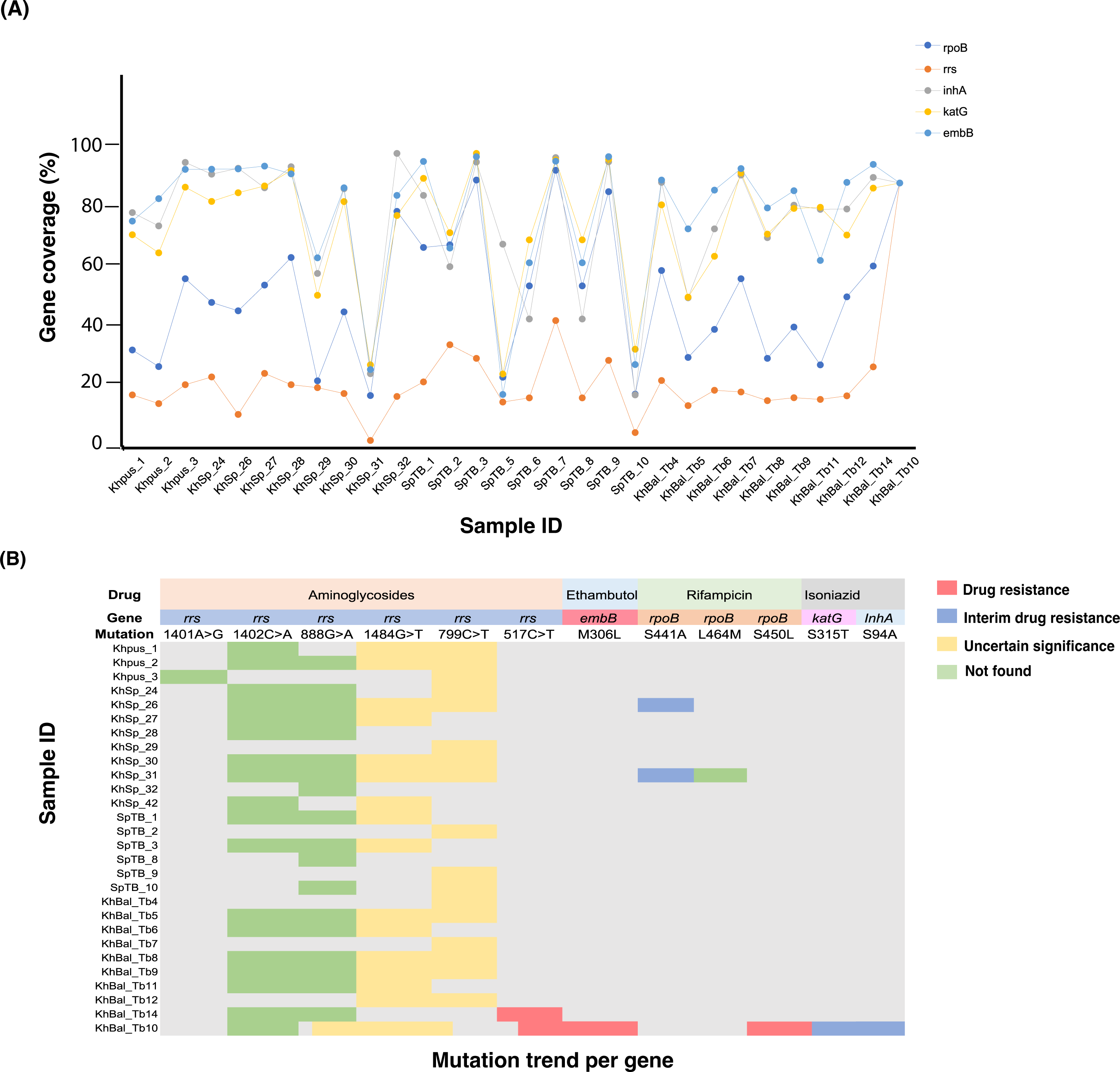
(A) Percentage coverage in resistance-associated genes: The percentage coverage of genes carrying mutations conferring drug resistance detected in twenty-one samples. The coverage trend per gene was similar in most of the isolates, with the *rrs* gene having the lowest and *embB* having the highest coverage reads mapping to these genes was detected in 99.9% of the samples with median percentage coverage 85% (IQR= 61%-94%) **Mutation trend in drug resistance genes (B):** Majority of the samples had mutation in *rrs* gene causing resistance to aminoglycosides. However, most of the mutations were classified as mutations of uncertain significance (yellow) as per WHO or not found in the existing catalogue (green). Samples with multiple mutations causing resistance to more than one drug were also observed.

Mutations associated with ethambutol, streptomycin, ethionamide and aminoglycosides were also detected in the same sample. (**Figure 3B**). This sample was classified as pre-XDR as per our WGS analysis. We detected a non-synonymous mutation in *rpoB* gene (Ser441Ala) in two samples which were classified as RIF sensitive by Xpert MTB/RIF. However, Ser441Ala *rpoB* mutation has been classified as an important mutation causing RIF interim resistance as per the WHO catalog.

### Phylogenetic analysis and mixed infection detection

Out of the 30 samples, phylogenetic analysis successfully assigned lineages and sub-lineages to 21 samples (70%). This included BAL (n=8), pus (n=3) and sputum (n=10) samples. The predominant lineage among these samples was lineage 1 (Indo-oceanic), which was detected in 13 out of 21 samples (61.9%). These findings are in concordance with the previously reported prevalence of *Mtb* lineages in India. Indo Oceanic Lineage is one of the major widespread lineages prevalent in specific geographical regions and are known to have high mortality rate (35). Other lineages that were identified included lineage 2, 3, 4, 6, 7, and 9 (**Table 3**).

**Table 3:** List of *M.tb* lineages in various samples predicted on *TBProfiler* using WGS data.

| Samples | Sample Types | Lineage |
| --- | --- | --- |
| KHSpTB24 | Sputum | Lineage 1, Lineage 1.1.1 |
| KHPus_TB1 | Pus | Lineage 1.1.1 |
| SpTB3 | Sputum | Lineage 1.2.1.2(indo-oceanic) |
| KHBal_TB10 | BAL | Lineage 1.2.2.2(indo-oceanic) |
| KHPus_TB2 | Pus | Lineage 2.2.1,Lineage 4.6.3, Lineage 6.3.1, Lineage 4.6.2.1 |
| KHSpTB30 | Sputum | Lineage 6.3.1 |
| KHSpTB28 | Sputum | Lineage_1 |
| KHSpTB29 | Sputum | Lineage1 |
| KHSpTB31 | Sputum | Lineage1,Lineage4.9 |
| KHSpTB26 | Sputum | Lineage1.1.1 |
| KHSpTB27 | Sputum | Lineage1.1.1 |
| KHBal_TB8 | BAL | Lineage4.9 |
| SpTB5 | Sputum | Lineage4.9 |
| SpTB1 | Sputum | Lineage4.9,Lineage 1.2.2.2 |
| KHPus_TB3 | Pus | Lineage6.3.1,Lineage4.3.1.1,Lineage1.2.1.2.1 |
| KHBal_TB4 | BAL | Lineage7 and Lineage9 |
| KHBal_TB5 | BAL | Lineage7 and Lineage9 |
| KHBal_TB11 | BAL | Lineage1.2.1.2, 1.1.1.1 |
| KHBal_TB12 | BAL | Lineage6.3.1, lineage3.1.2.2 |
| KHBAL_TB14 | BAL | Lineage6.3.1, lineage3.1.2.2 |
| KHBal_TB6 | BAL | Lineage 1, 2.2.1, 4.6.2.1, 1.2.1.2.1 |
| KHBal_TB9 | BAL | Lineage1.2.1.2 |

We observed a high frequency of mixed infection in BAL and pus samples when compared with sputum. Overall, mixed infections were detected in eight samples (BAL=4/8, Pus=2/3 and Sputum= 2/10). KHPus_TB2 and KHPus_TB3 were infected with four and three different strains respectively. These included East Asian, Euro-American, West African II, Euro-American and Indo-Oceanic lineages. Higher background and low sample quality of sputum samples may explain the non-detection of strains with low copy number. These findings are in line with higher median percentage genome coverage of BAL (median = 15.4%) and pus (median =19.6%) samples than sputum (10.5%) despite low bacilli count as per smear microscopy.

## Discussion

Due to significant advancement in the whole genome sequencing technologies in recent years, there has been an increase in accessibility to cost-effective sequencing platforms. However, the majority of sequencing studies use *Mtb* DNA extracted from cultures, which takes several weeks to months and requires BSL-3 facility. Moreover, WGS sequencing from cultures may create a bias due to DNA extracted from selected colonies and may not detect mixed infections (8, 36). Direct WGS from clinical samples could be a solution to eliminate these shortcomings; however, without any enrichment, it provides poor coverage that cannot be used for further analysis. To address these gaps, we used ligand-based magnetic beads to enrich *Mtb* from clinical samples and performed WGS. To our knowledge, this is the first study demonstrating the use of magnetic beads to enrich *Mtb* from clinical samples for WGS and direct sequencing of smear negative and EPTB samples .We demonstrated that concentrating *Mtb* prior to DNA extraction significantly increased the sequencing positivity in the smear negative samples. We were able to detect *Mtb*-specific reads in the majority of the samples 88.2% of the samples, with 35.3% smear negative. In addition to confirming the drug resistance detected in routine diagnostics, we also identified mutations that confer resistance to other anti-TB drugs. These mutations are generally not covered by PCR-based tests due to limitations in the number of targets that can be screened. Direct WGS sequencing also allowed detection of mixed infections and classification of *Mtb* into respective lineages and sub lineages in the majority of the samples (70%).

Studies have shown that performing direct sequencing of *Mtb* from clinical samples without any enrichment does not yield enough data to carry out the basic genomic analysis. In a study by Doughty and colleagues, eight AFB^3+^ sputum samples were processed without any enrichment followed by WGS (17). Despite using samples with high bacillary load, the maximum *Mtb* genome coverage depth obtained was 0.7X. Seven of eight samples could be assigned to the *M. tuberculosis* complex, but none had sufficient data for drug susceptibility prediction. Using magnetic bead enrichment, we obtained upto 18.7x coverage for AFB^3+^ sample and were able to achieve sufficient coverage to predict drug resistance and mixed infections. In addition to sequencing *Mtb* enriched from sputum, we also sequenced *Mtb* from BAL and pus samples for which no direct sequencing data have been available thus far. It is interesting to note that pus and BAL samples which were negative on smear microscopy had relatively higher coverage than the smear positive sputum samples. This could be explained due high viscosity of sputum samples that might lead to trapping *Mtb* in the sputum inhibiting binding to the beads. An additional sputum thinning step may further improve the yield. Also, presence of other bacteria which would potentially bind to the beads and which cause saturation may also affect the yield. Further studies on a larger sample size are needed to confirm these findings.

The World Health Organization (WHO) has emphasized on prioritizing the affordable and accessible point-of-care TB diagnostics, including for DST. As the WHO now recommends sequencing-based assays for *M.tb* drug resistance determinants, the relevant equipment and expertise may be increasingly available in public health laboratories in tuberculosis-endemic settings. (1). Currently available molecular assays target markers for a limited number of drugs and are not usually designed to incorporate a growing list of resistance mutations for additional drugs (37). These assays can sometimes lead to false negative results or underrepresentation of mutations in a population. Although diagnostics such as the Fluorotype® MTB test (Hain, Germany) and the Anyplex™ II MTB/MDR can detect resistance to fluoroquinolones and second-line injectable drugs, their sensitivity ranges from 69.1% to 99.2% (38–40). In a study to evaluate *Mtb* clinical isolates from Congo it was found that there was a misidentification of fluoroquinolone resistance by line probe assay due to a double substitution T80A-A90G in *gyrA* gene (41). Furthermore, additional assays are currently needed for surveillance or outbreak detection adding to the overall cost. Therefore, developing a method that allows large-scale screening of mutations directly from clinical samples is an attractive prospect. We compared the WGS data obtained from our study with a comprehensive list of mutations published by the WHO that are either associated with or potentially linked to resistance against anti-TB drugs. In addition to confirming mutations detected on Xpert MTB/RIF, we detected a non-synonymous Ser441Ala mutation in *rpoB* gene in two samples not covered by routine diagnostic assays. This mutation has been classified as a mutation causing interim resistance to rifampicin. Individuals carrying such mutations can benefit from dose adjustments. One such example is patients with mutations in *katG* and *inhA* genes. Mutations in *katG* confer high-level isoniazid resistance, whereas mutations in *inhA* causes low-level resistance to isoniazid (42). Studies have shown that frequency of these mutations varies geographically and only 10% of the population have both *katG* and *inhA* mutations (43). Therefore, patients with only *inhA* mutation may benefit from high doses of isoniazid (44). Identifying the resistance and the frequency patterns of such mutations may help in making decisions regarding drug regimen.

Another relevant application of direct whole-genome sequencing is the study of *Mtb* genetic diversity in sputum samples, which might better reflect the within-patient bacterial populations. So far, the extent to which culture reduces the actual *Mtb* genetic diversity remains unclear. It is also likely that additional diversity can be identified in settings where several lineages coexist and generate mixed infections. Studies have reported cases of reinfection by a second *Mtb* strain and occasional infection with more than one strain making it difficult to assign the accurate treatment regimen (11, 12). Infection with multiple strains of *Mtb* can complicate the interpretation of drug susceptibility testing (DST) results and the detection of epidemiological links (13). Using direct sequencing on enriched samples, we detected mixed infection (26.6%) of the samples. It is interesting to note that infection with upto four strains of *Mtb* was detected in pus samples. Studies have shown that patients harboring multiple strains of *Mtb* have poor treatment outcomes (45).

The findings of our study are subject to several limitations. Firstly, we only sequenced samples enriched with magnetic beads and did not have whole genome sequencing data on samples without enrichment to quantify the loss of *Mtb* cells during DNA extraction.

Furthermore, while we observed a significant difference in terms of percentage reads mapping to H37Rv when compared between sputum, BAL, and pus, we analyzed a limited number of pus samples (n=4) and further studies on a larger sample size would be required to validate these findings. We did not perform a culture DST comparison of the resistance data for the confirmation, although the resistant data is compared with the WHO mutation catalogue. Additionally, we only analyzed three AFB3+ samples to demonstrate that a magnetic bead-based method can be used to enrich *Mtb* for WGS to achieve sufficient coverage for basic analysis. While coverage obtained for most of the samples in our study was better than previously reported study and allowed the basic analysis, this technique still requires further optimization to perform complex whole genome sequencing analysis that require higher coverage.

## Conclusions

Amid the growing evidence that whole genome sequencing of *Mtb* can detect drug resistance, predict pathogen prevalence and evolution patterns, and identify mixed and co-infections, there is a need for platforms that enable the direct whole genome sequencing of *Mtb* from clinical samples eliminating bias. We used magnetic bead enrichment method to capture *Mtb* prior DNA extraction to perform whole genome sequencing. Our strategy can be used to perform WGS on smear negative and difficult to culture extra pulmonary TB samples. The enriched samples can also be used to perform targeted genome sequencing to achieve higher coverage. This strategy would be especially useful in samples which are not detected by routine diagnostic assays due to low bacillary load. Magnetic bead enrichment is easier to perform, is cost-effective and can be scalable for use in settings where TB burden is greatest.

## Data availability

Data supporting the findings of this manuscript have been submitted to NCBI and can be accessed through the SRA submission portal with accession number PRJNA1052384 (https://submit.ncbi.nlm.nih.gov/subs/sra/SUB14001592/overview).

## Authors contribution

### Declaration of competing interest

The authors declare that they have no known competing financial interests or personal relationships that could have appeared to influence the work reported in this paper.

## Acknowledgements

Authors would like to acknowledge all the technical staff and patients from Manipal Hospital for their cooperation during the study.

## Funding

RV is supported by Ramalingaswami Fellowship, Department of Biotechnology (DBT) Govt. of India (ID No. BT/HRD/35/02/2006)

---

**Supplementary figure 1:**
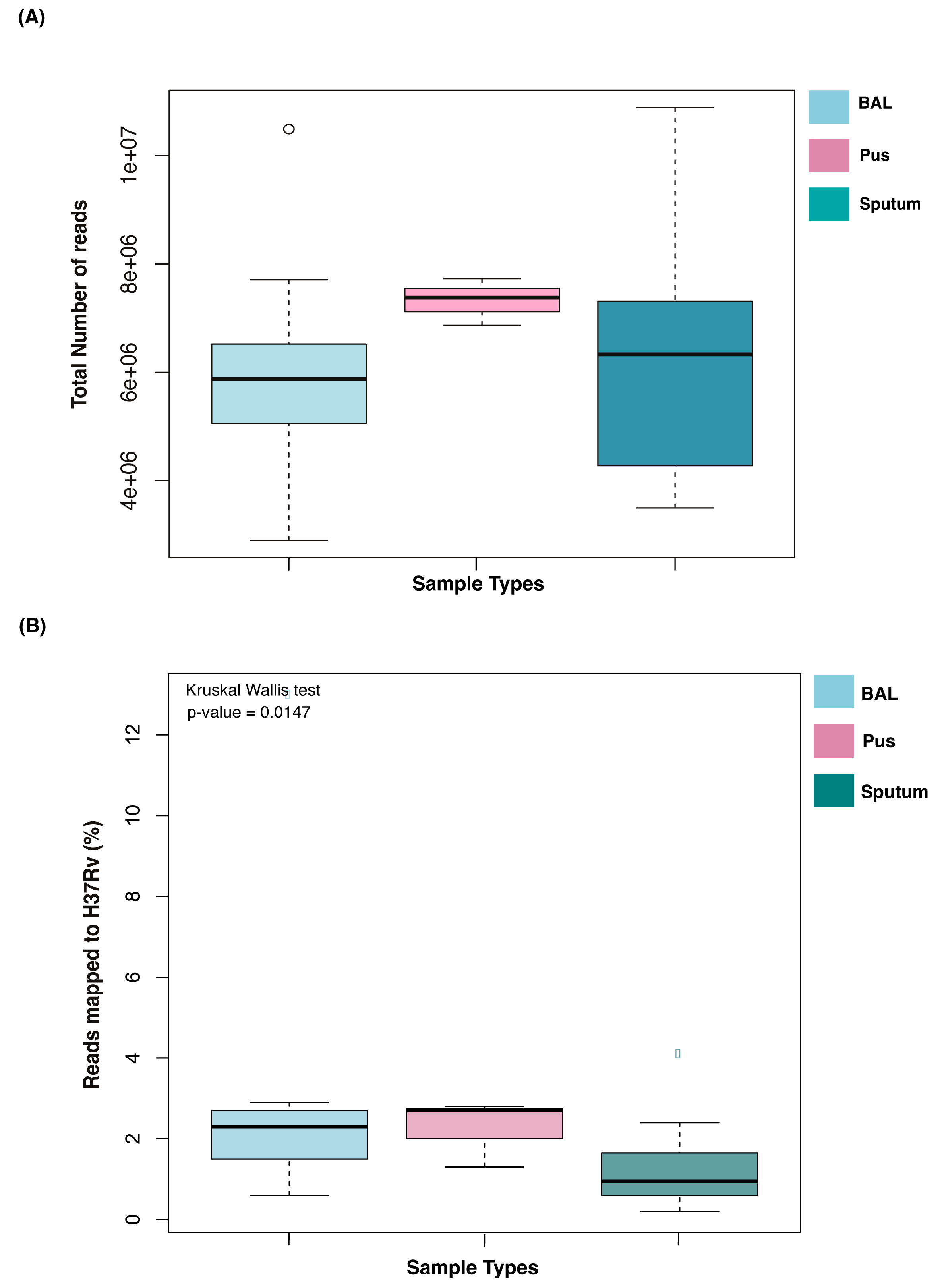
**(A)** Total number of reads identified in various sample types (BAL, Pus and sputum) after quality filter. **(B)** Percentage of reads mapped to H37Rv genome.

## Notes

### Competing Interest Statement

The authors have declared no competing interest.

### Author Declarations

The study was approved by the institutional review board of Kasturba medical college, Manipal Academy of Higher Education (IEC:526/2018).

